# The trans-kingdom spectrum of Mpox-like lesion pustules of suspect patients in the Mpox subclade Ib outbreak in Eastern Democratic Republic of the Congo

**DOI:** 10.1101/2025.01.04.24319669

**Authors:** Leandre Murhula Masirika, Benjamin Hewins, Ali Toloue Ostadgavahi, Mansi Dutt, Léandre Mutimbwa Mambo, Jean Claude Udahemuka, Pacifique Ndishymie, Justin Bengehya Mbiribindi, Freddy Belesi Siangoli, Patricia Kelvin, Morgan G. I. Langille, David J Kelvin, Luis Flores, Gustavo Sganzerla Martinez, Anuj Kumar

## Abstract

During infectious disease outbreaks, acquiring genetic data across various kingdoms offers essential information to tailor precise treatment methodologies and bolster clinical, epidemiological, and public health awareness. Metagenomics sequencing has paved the way for personalized treatment approaches and streamlined the monitoring process for both co-infections and opportunistic infections. In this study, we conducted long-read metagenomic DNA sequencing on Mpox-like lesion pustules in six suspected patients during the MPXV subclade Ib outbreak in Eastern Democratic Republic of the Congo. The sequenced data was taxonomically classified per the bacterial, fungal, and viral composition. Our results show a wide spectrum of microorganisms present in the lesions. Bacteria such as *Corynebacterium amycolatum, Gardnerella vaginalis, Enterococcus faecium, Enterobacter clocae, Staphylococcus epidermidis*, and *Stenotrophomonas maltophilia* were found in the lesions. The viral classification of the reads pointed out the absolute predominance of Monkeypox virus. Taken together, the outcomes of this investigation underscore the potential involvement of microorganisms in mpox lesions and the potential role that co-infections play in exacerbating disease severity and transmission during the MPXV subclade Ib outbreak.

## INTRODUCTION

Monkeypox virus (MPXV) is the causative agent of mpox and is endemic in regions of Central and East Africa, including the Democratic Republic of the Congo (DRC). The first human Mpox infection occurred in 1970 in DRC and since then sporadic outbreaks have increased in frequency [1]. In September 2023, sustained human-to-human transmission of the newly identified MPXV subclade Ib virus in eastern DRC led the World Health Organization to declare a Public Health Emergency of International Concern [2-4] as travel-related cases were observed in Europe, Asia, and North America, marking the first time MPXV Clade I viruses were recorded outside the African continent. The outbreak exhibited transmission dynamics not previously observed during other Clade I mpox outbreaks, such as heterosexual transmission among professional sex workers (PSWs) and sustained (nonsexual) community-level transmission in adults, adolescents, and children [5]. Symptoms observed during this outbreak ranged from mild to severe and were characterized as a febrile illness with a progressive rash (macules, papules, vesicles, pustules, and scabbing) starting in the hands/feet, or genitals, and spreading over the body.

A detailed picture of the microbial makeup of mpox lesions during the subclad Ib outbreak is necessary to better understand why, specifically, this novel Clade I virus is so efficiently transmitted between humans, a characteristic rarely documented in Clade I mpox [6,7]. In much of Africa, mpox vaccines (Jynneos®, Imvamune®) are not readily available. Many available doses were preferentially procured and distributed among high-income countries during the 2022 global mpox Clade IIb outbreak, placing many endemic regions in Africa at risk of ongoing and worsening outbreaks. Furthermore, the lack of routine screening of sexually transmitted illnesses (STI) in low- and middle-income countries (LMICs) in Africa, such as DRC, is virtually nonexistent [8]. As transmission of subclade Ib appears to be occurring via heterosexual contact (and community spread), the lack of STI screening exacerbates transmission. Compounding the previous issues in these settings is the lack of infrastructure, reagents and supplies, and trained personnel to perform testing/screening.

The role of co-infection and super-infection during mpox infection is well-documented. For example, a recent systematic review determined that of 6345 confirmed mpox cases, 40.32% of individuals (N= 2,558) also had human immunodeficiency virus (HIV) co-infection [9]. The true number of HIV-mpox co-infected individuals in LMICs in Africa likely exceeds 40.32%, given that the majority of study locations reported in the review occurred in high-income countries, where contraception, STI screening, and access to healthcare is much greater. In addition to reduced immune system function caused by HIV, and thus the ability to stave off additional diseases, patients with Mpox are often diagnosed with other infections. These can include STIs (i.e., syphilis, chlamydia, and gonorrhea), herpes simplex virus (HSV), varicella zoster virus (VZV), cytomegalovirus, and species of *Mycoplasma* [10-16]. Given the morphological overlap of mpox, HSV, and VSV, broad STI screening and PCR should be incorporated as standard of care during suspected cases of mpox, where resources permit. Although genital mpox lesions in sexually active individuals are explained by sexual transmission, transmission during the subclade Ib outbreak is complicated by cases of community spread, where contact with infected materials (fomites) has been shown to spread the disease.

Currently little is known regarding the viral, bacterial, and fungal communities present in mpox-associated lesions. A greater understanding of these lesions may provide additional insights into the transmissibility, genetic diversity, and host specificity of subclade Ib. To address these gaps, we hypothesize that there is a wide spectrum of pathogens present in lesions caused by mpox, likely functioning synergistically to exacerbate disease state and transmission. To test our hypothesis, we investigated the trans-kingdom spectrum of sequenced lesion samples to ascertain their viral, bacterial, and fungal composition.

## METHODS

### Study design and sample collection

Samples were collected from lesion swabs obtained from suspected cases of mpox infection from six patients admitted to the Kamituga General Hospital in the city of Kamituga, South Kivu, Democratic Republic of the Congo. The DNA extraction was performed using the Qiagen DNeasy Blood & Tissue Kit (Qiagen, Hilden, Germany) according to the manufacturer’s instructions. The quality and quantity of DNA was determined using a Qubit 4 Fluorometer with the Qubit 1X dsDNA HS Assay Kit (Thermo Fisher Scientific, Waltham, MA, USA). The test was performed using a 1 µl sample volume according to the manufacturer’s protocol. For the successful preparation of libraries, the use of this high-sensitivity assay was crucial for accurate DNA quantification, even at low concentrations.

### Metagenomics Sequencing

The sequencing libraries were prepared with two distinct kits: for R10 flow cells, the Rapid Sequencing DNA - PCR Barcoding Kit 24 V14 (SQK-RPB114.24), and for R9 flow cells, the Rapid PCR Barcoding Kit 24 (Version 14, SQK-RPB004), both products of Oxford Nanopore Technologies (ONT). The procedure involved several steps including initial DNA tagmentation, barcode addition using rapid barcode primers, PCR amplification, and library cleanup with AMPure XP magnetic beads (Beckman Coulter), followed by a final quantification step with the Qubit 4 fluorometer. The libraries were then loaded on the R9 or R10 flow cells using proper flow cell priming and library loading procedures per ONT protocols. Sequencing runs were extended to 24 hours with real-time base calling enabled through MinKNOW software (version 23.11.5, Oxford Nanopore Technologies) for live monitoring and data quality assessment. Nanopore raw reads were base called using the MinKNOW software (ONT, version 23.11.5) with a minimum Q score of 8.0.

## Taxonomic Annotation and Visualization

Nanopore reads had their taxonomic composition annotated Kraken2 (version 2.1.3) [17] using the PlusPF database, composed of RefSeq archaea, bacterial, viral, protozoa, and fungi (1/12/2024). The Kraken2 outputs were converted to a report file, which was carried over, read, and parsed in Python (version 3.12) to plot. In addition, the Kraken2 reports were visualized using the R package Pavian (version 1.2.1 running on R version 4.1.2).

## RESULTS

### Metagenomics sequencing

Lesion swabs from six Mpox suspect cases were obtained and sequenced using ONT. In Figure 1, we show a patient, representative of the subclade Ib outbreak in Eastern DRC, exhibiting multifocal, erythematous maculopapular mpox-like lesions distributed extensively across the left upper extremity, thoracic region, dorsal aspect, cervical region, and facial integument. The number of raw reads varied from 964K to 3M. When considering reads that passed the minimum Q score of 8, the basecalling process generated from 3.18 to 10.45 giga bases. In addition, the raw reads were classified using Kraken2 to determine their metagenomic composition of viruses, bacteria, archaea, and fungi (Table 1).

**Table 1.** Metagenomic sequencing and taxonomic annotation of six mpox lesion samples.

| Sample | Sequencing |  |  | DNA-to-DNA (Kraken2) |  |  |
| --- | --- | --- | --- | --- | --- | --- |
|  | Raw reads (million) | Total bases (Gb) | Passed bases (Gb)<sup>1</sup> | Bacteria/archaea | Fungi | Viruses |
| 1 | 0.964 | 3.18 | 2.43 | 5,009 | 25 | 17,896 |
| 2 | 2.07 | 7.02 | 6.33 | 17,800 | 194 | 61,808 |
| 3 | 3 | 11.18 | 10.45 | 74 | 14 | 53,868 |
| 4 | 1.29 | 5.13 | 4.59 | 874 | 15 | 87,600 |
| 5 | 1.96 | 4.64 | 3.80 | 13 | 1 | 6,821 |
| 6 | 1.58 | 5.23 | 4.63 | 2,820 | 11 | 41,620 |

<sup>1</sup>Q score $\geq 8$ .

**Figure 1.**
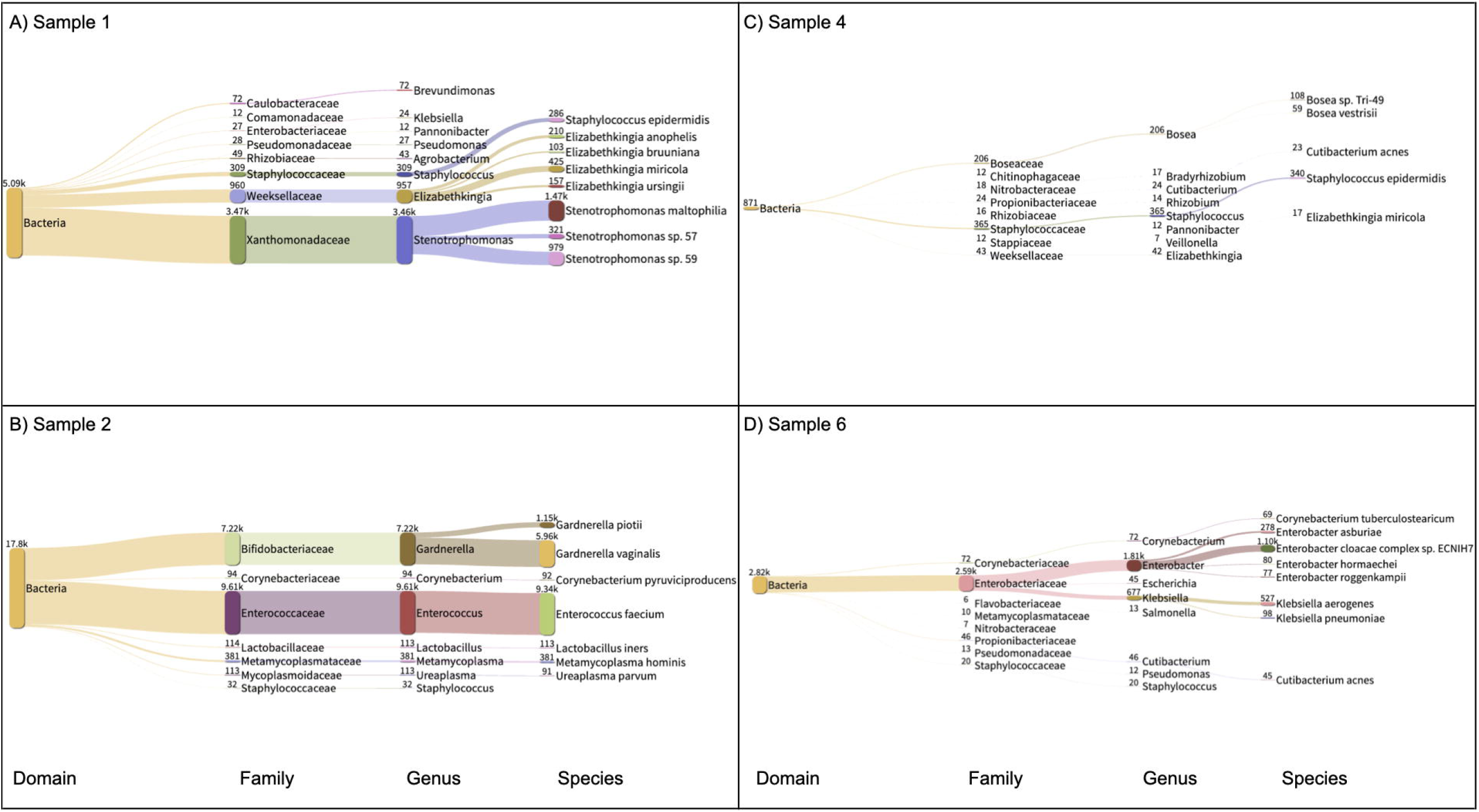
DNA to DNA bacterial taxa classification of mpox lesion samples. We isolated the bacterial domain of the Kraken2 taxonomy classification of Sample 1 (Figure 1-A), Sample 2 (Figure 1-B), Sample 4 (Figure 1-C), and Sample 6 (Figure 1-D). The thickness of the horizontal links between taxonomical ranks represents the number of reads. The plots were generated with Pavian (version 1.0, running on R version 4.1.2). The taxonomical ranks displayed are Domain, Family, Genus, and Species. The numbers shown in superscript on the left side of each taxonomical rank represent the total number of reads whose DNA was classified.

### Trans-kingdom taxonomic annotation

First, we analyzed the bacterial DNA classified in each sample (Figure 2). Samples 3 and 5 had 74 and 13 reads classified as bacteria, respectively; thus we opted to leave them out of Figure 2 due to low bacterial reads. On the other hand, samples 1, 2, 4, and 6 had 5.09K, 17.8K, 871, and 2.82K DNA reads classified as belonging to the bacterial domain. No bacterium was found in common across all four samples. *Staphylococcus epidermidis* was found in samples 1 and 4 with 286 and 340 reads, respectively. Moreover, in sample 1, a total of 3.46K reads had their DNA classified as belonging to the Stenotrophomonas genus (*S. maltophilia, S. sp*. 57, and *S. sp*. 59). Sample 2 had 5.96K reads of *Gardnerella vaginalis* and 9.34K of *Enterococcus faecium*. Sample 6 had 1.81K reads classified in the Enterobacter genus with 1.10K reads belonging to the bacterium *Enterobacter cloacae complex sp*. ECNIH7. We also explored the DNA-to-DNA taxonomy classification of the six samples across fungi. In sample 2, which had 194 reads classified as fungus, 184 reads were classified as belonging to the DNA of *Nakaseomyces glabratus*. The remaining fungal reads classified in the other samples were not mapped to any specific organism. Finally, we explored the DNA-to-DNA taxonomy classification of the six samples across viral species (Figure 3). The number of DNA reads classified as MPXV reads was above 99% of the total viral reads across the six samples.

**Figure 2.**
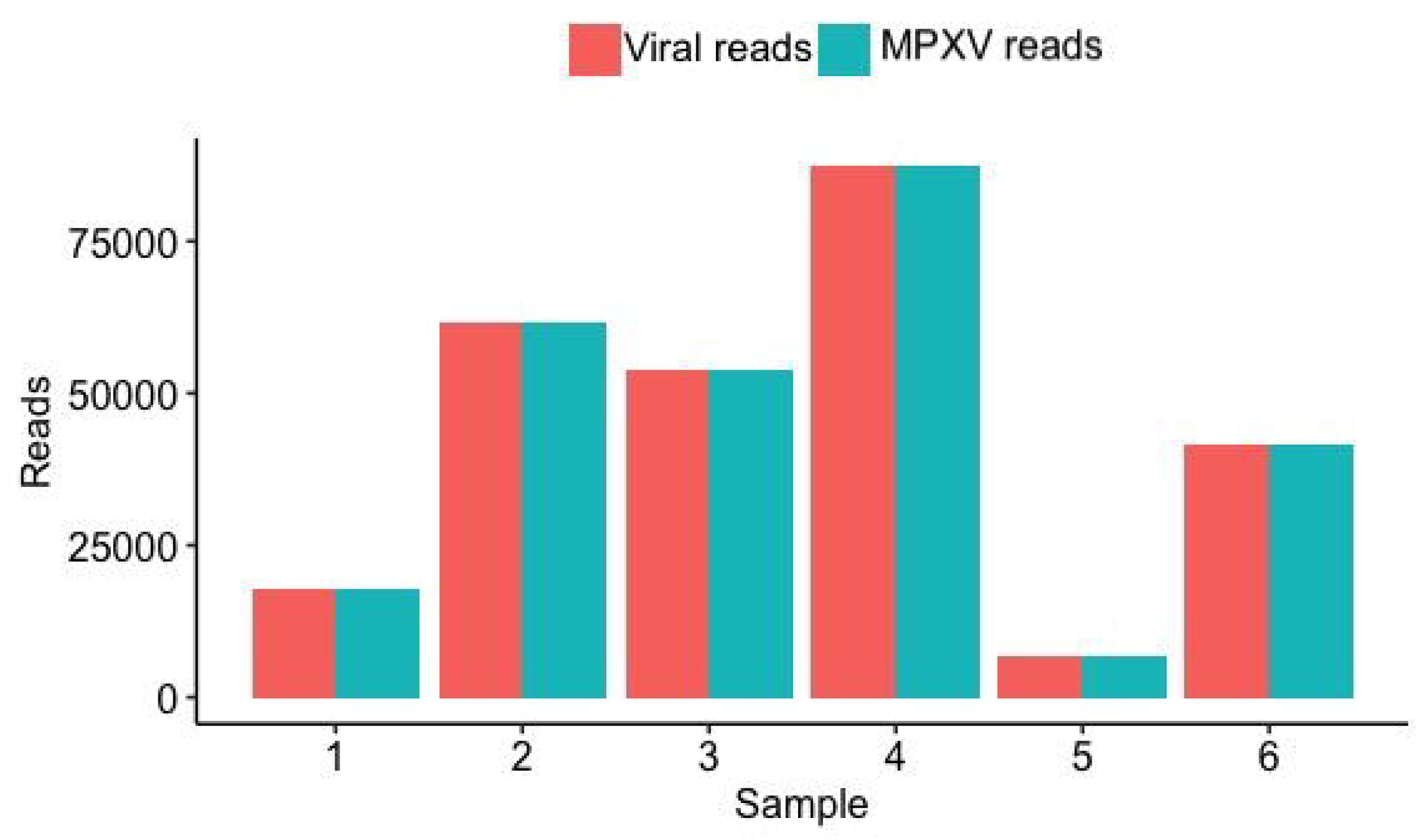
The taxonomy distribution of viral DNA reads. We show in the x-axis each sample and in the y-axis the absolute number of viral and MPXV reads classified in a DNA-to-DNA approach using Kraken2.

## DISCUSSION

In our study, we determined the trans-kingdom spectrum of microorganisms found in pustule lesions of mpox-suspect patients with metagenomic sequencing and taxonomical classification. Each sample presented in this study provided reads classified as viruses, bacteria, and fungi, which all might play important roles in contributing to pathogenicity.

Beyond causing skin rash and lesions, Mpox can trigger a strong effect on the host immune response, characterized by impaired Natural Killer cell function, lymphopenia, increased antibodies, increased blood monocytes and granulocytes, immune evasion, cytokine storm, inhibition of the host complement system, and antibody-dependent enhancement [18]. Therefore, during Mpox infection, infected individuals may be more prone to bacterial, viral, and fungal infections as the immune response is undergoing several rapid alterations in response to the initial pathogenic challenge (MPXV) [19]. We argue that initial infection with Mpox may have resulted in an immunocompromised state, facilitating further co-infection with the bacteria identified in our analysis.

Before the widespread emergence of next generation sequencing (NGS) technologies, the identification and interaction of microbes relied largely on in vitro culturing. Only within the last decade have the cost and barriers associated with NGS and metagenomic profiling decreased, allowing greater integration into clinical diagnostic practice [20,21]. Rather than isolating and culturing a single pathogen, metagenomic profiling provides insights into the viral, bacterial, and fungal makeup of a patient sample. The specificity of metagenomic profiling reduces false positives and provides a more sensitive, unbiased delineation of the causative pathogenic agents responsible for a given infection. Clinical metagenomics may also be leveraged to provide a tailored approach for treating a patient-by-patient disease spectrum, where co-infections often complicate the initial diagnosis [22].

In terms of viral classification, MPXV-classified reads were predominant across the six samples analyzed in this study. No other virus was found in any other sample. A low number of reads were classified as fungi. On the other hand, bacteria were found across samples. Despite not identifying a common bacterium in all samples, we observed samples 1 and 4 having reads classified as *S. epipermidis*. there is evidence of this organism having evolved to not infect humans [23] thus configuring a commensal relationship. A total of 3,460 reads of sample 1were classified as pertaining to the *Stenotrophomonas* genus (with predominance of the bacterium *S. maltophilia*). skin infections of *S. maltophilia* have been classified as very rare among non-immunocompromised patients [24], which might suggest the patient acquired the infection when their immune system was already stressed. In addition our taxonomy classification confirmed the presence of *E. faecium* and *G. vaginalis* in sample 2. Both microorganisms have been reported to cause infection in humans [25, 26]. In sample 6, we identified bacterial reads classified to the *Enterobacter* genus, with more reads classified as the species *E. cloacae*. Despite the lack of evidence of direct diseases caused by *E. clocae*, this pathogen is commonly found in nosocomical infections, with a high-rate of mortality associated with the *Enterobacter* genus [27, 28].

Limitations of our study include the possible sample bias and lack of sequence information for control skin samples as comparators for the mpox pustules. Moreover, the general lack of health infrastructure in the South Kivu province limited testing/sampling capacity, and ongoing armed conflict in the region reduced our ability to test for HIV, potentially introducing cofounders and influencing the immune status of study subjects. Finally, despite having identified opportunistic organisms, such as *S. maltophilia*, which generally causes infections in already immunocompromised individuals, the lack of longitudinal data of our study makes it difficult to speculate the trans-kingdom spectrum of patients prior to their MPXV infection.

In conclusion, our trans-kingdom analysis identified potential bacteria with known and unknown pathological functions coexisting within mpox virus skin lesions. These samples were collected during an ongoing MPXV outbreak in the South Kivu province of DRC. Findings from this study should serve as a warning sign for public health authorities that additional microorganisms may be contributing to the disease severity and transmissibility observed during the subclade Ib outbreak. Furthermore, our findings underscore the importance of continued genomic surveillance in mpox endemic regions and highlight the need for improved routine STI screening in these settings. Further investigation is needed to determine if human populations living in different geographical locations have similar or different flora in mpox lesions.

## Data Availability

All data produced in the present study are available upon reasonable request to the authors.

## Acknowledgements

The authors would like to thank Dr. Nikki Kelvin for editing the final draft of the manuscript. We thank the Digital Research Alliance of Canada which, through ACENET, provided in-kind computational resources for running the metagenomic analyses depicted in this study.

## Ethical Statement

Ethical clearance for conducting this study was approved by the Ethical Review Committee of the Catholic University of Bukavu (Number UCB/CIES/NC/022/2023). All study participants were introduced to the observational study and given the option to participate by providing informed consent or, in the case that the participant was a minor, parental permission or consent was obtained. In addition, we obtained written consent from the patient whose picture is shown in Figure 1.

